# BNT162b2 induces robust cross-variant SARS-CoV-2 immunity in children

**DOI:** 10.1101/2022.05.18.22275283

**Authors:** Yannic C Bartsch, Jessica W Chen, Jaewon Kang, Madeline D Burns, Kerri J St Denis, Maegan L Sheehan, Jameson P Davis, Alejandro B Balazs, Lael M Yonker, Galit Alter

## Abstract

Currently available mRNA vaccines are extremely safe and effective to prevent severe SARS-CoV-2 infections. However, the emergence of novel variants of concerns has highlighted the importance of high population-based vaccine rates to effectively suppress viral transmission and breakthrough infections. While initially left out from vaccine efforts, children have become one of the most affected age groups and are key targets to stop community and household spread. Antibodies are central for vaccine induced protection and emerging data points to the importance of additional Fc effector functions like opsononophagocytosis or cytotoxicity, particularly in the context of variants of concern that escape neutralizing antibodies. Here, we observed delayed induction and reduced magnitude of vaccine induced antibody titers in children 5-11 years receiving two doses of the age recommended 10 μg dose of the Pfizer SARS-CoV-2 BNT162b2 vaccine compared to adolescents (12-15 years) or adults receiving the 30 μg dose. Conversely, children mounted equivalent or more robust neutralization and opsonophagocytic functions at peak immunogenicity, pointing to a qualitatively more robust humoral functional response in children. Moreover, broad cross-variants of concern responses were observed across children, with enhanced IgM and parallel IgG cross-reactivity to variants of concern (VOCs) in children compared to adults. Collectively, these data argue that despite the lower magnitude of the BNT162b2 induced antibody response in children, vaccine induced immunity in children target VOCs broadly and exhibit enhanced functionality that may contribute to attenuation of disease.

## Introduction

Rapid vaccine developments and distributions have dramatically mitigated the disease burden of SARS-CoV-2. Children and adolescents, who were mostly spared from the initial surge of COVID-19, were excluded from early vaccine efforts. However, as the pandemic continued, it has become evident that children, too, can suffer from severe COVID-19^1^ as well as long-term disease^2^. There has been a marked increase in SARS-CoV-2 cases in children under the age of 18^3^, potentially fueled by loosened mask mandates^4^ and the widespread transmission of novel variants of concern (VOC) including B.1.1.529 (Omicron) and its sub-variants^5^. In some areas in the US with low vaccination rates and lack of herd immunity, there is a four-fold increase of hospitalizations in children compared to areas with high vaccine rates^6^. Additionally, children are at risk of developing the severe, post-COVID-19 illness, multi-inflammatory syndrome (MIS-C), weeks after they were exposed to the virus pointing to the importance of vaccine availability to all ages^7^. Moreover, increasing numbers of cases of long-COVID have begun to accrue in children, with unexpected symptoms including brain fog/dizziness, hair loss, stuttering or palpitations^8^. Meanwhile, mRNA vaccines are available through emergency use authorization (EUA) for children five years and older, however, due to safety and tolerance concerns, mRNA vaccines are available with an adjusted lower dose for children under 12 years of age. It is unclear whether this dose adjustment will negatively impact immunogenicity and lead to a more variable outcome^9^. Moreover, the early unexperienced immune system matures over the first decades of life and humoral responses to vaccines including mRNA vaccines can differ considerably to those observed in adults^10^.

Antibodies against SARS-CoV-2 Spike are pivotal correlates of protection from infection and severe COVID-19^11^. Currently available mRNA vaccines are able to induce comparably high Spike specific binding titers across different emerging VOCs^12^. While antibody-mediated viral neutralization is often used as marker of protection, compared to binding, neutralization is more sensitive to viral evolution in emerging VOCs^13^. Along these lines, reduction of *in vitro* neutralization^14,15^ is not reflected in loss of vaccine protection from severe disease in real-life^16,17^, and other antibody mediated effector functions including opsonophagocytosis, complement activation, and NK cell cytotoxicity, which have less epitope restriction, might be equally important to block transmission and disease^18,19^. While SARS-CoV-2 vaccines are able to induce broad binding and functional antibodies in adults, whether these responses also emerge in children, particularly in the setting of lower doses in younger children, remains unclear.

Here, to begin to define whether SARS-CoV-2 mRNA vaccines, and particularly the most widely distributed BNT162b2 vaccine, induce functional humoral immune responses in children, we comprehensively profiled vaccine induced immune responses in 32 children (5-11 years) receiving two doses of BNT162b2 with the age-specific recommended 10μg dose compared to adolescents (12-15 years, n= 30) and adults (16+ years, n=9) receiving the adult 30ug dose. Despite robust induction of SARS-CoV-2 Spike specific antibodies, we observed differences in the humoral immune responses across the groups, marked by slower induction and lower antibody titers in the children, but the induction of more robust functionality. While lower in magnitude, VOC-specific breadth was comparable across the groups, albeit effector functions to omicron were lowest in the 10 μg dosed children. Collectively, our data point to age and dose related differences in BNT162b2 induced humoral immunity, that may underly differences in viral breakthrough in the pediatric populations^20^ but point to the robust induction of functional humoral immune responses that may confer protection against severe disease across VOCs.

## Results

The Pfizer/BioNTech BNT162b2 SARS-CoV-2 vaccine has been approved under emergency use authorization in children 12-15 and 5-11 years old. While all individuals over 12 years of age are immunized with two doses of 30 μg BNT162b2, a two-dose regimen of a three-fold lower 10 μg dose was developed for children 5-11 years old. However, whether this dose adjustment and overall age-specific differences in immunity lead to differences in the humoral immune responses to SARS-CoV-2 mRNA vaccines remains unclear. To begin to investigate whether age and dose-dependent differences between the different age groups exists, we compared the humoral immune response in children 5-11 years old (n= 32, median age 9 years, 34 % female) receiving the age recommended 10 μg dose to adolescent 12-15 years old (n= 30, median age 13 years, 63 % female) or adults (n = 9, median age 19 years, 60 % female) receiving the 30 μg dose BNT162b2. Plasma samples were collected before the first (V0) and second dose (V1) as well as 2-4 weeks after the second dose (V2).

### BNT162b2 vaccination elicits robust SARS-CoV-2 specific isotype titers across the different age groups

To compare the humoral immune responses, we analyzed Spike and Receptor binding domain (RBD) specific IgM, IgA and IgG1 isotype titers (**Figure 1**). All individuals 12 years and older vaccinated with the 30 μg dose seroconverted (marked by an increase in titer) after the first dose. Likewise, most younger individuals immunized with 10 μg BNT162b2 seroconverted after the first dose (V1). Overall, IgM and IgA Spike and RBD titers tended to be highest in adults, whereas IgG1 titers were higher in the 30 μg vaccinated adolescent group. Furthermore, after one dose, Spike and RBD-specific IgG1 titers were significantly lower in children compared to adolescents (p=0.03) and adults (p<0.01) and titers remained lower in young children after the second dose. Interestingly, we noted that some individuals had detectable, albeit lower, Spike and RBD specific IgA and IgG titers at the pre-vaccination (V0) timepoint. None of the participants reported a known previous SARS-CoV-2 infection and we could not find serological evidence of pre-exposure marked by Nucleocapsid specific IgG1 (**Fig S1**). However, these pre-vaccine titers were highly correlated to Spike specific antibodies against the beta-coronavirus OC43 and HKU1, pointing to cross-coronavirus immunity. Importantly, the frequency of these responses did not differ across the age groups but they did correlate with the magnitude of the vaccine induced responses after the 1^st^ or 2^nd^ dose of the vaccine (**Fig S2**). Collectively, these data argues that BNT162b2 vaccination can induce robust SARS-CoV-2 specific antibody responses in adolescent and adults but result in a more variable response in children 5-11 years old receiving the three-fold lower dose.

**Figure 1.**
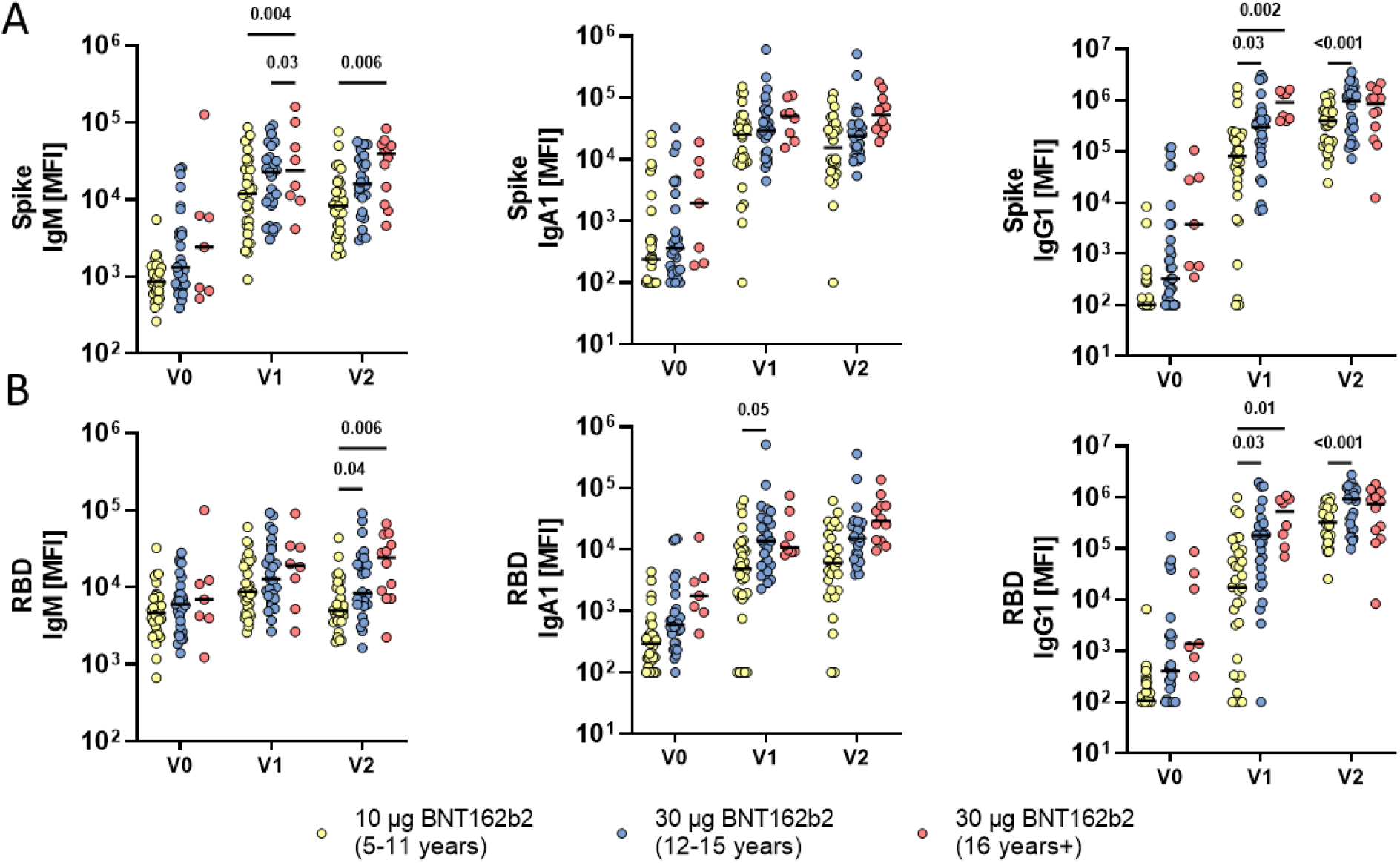
Vaccination with BNT162b2 mRNA in children results in antibody class switching and produces robust immune response. Relative SARS-CoV-2 wild-type (A) spike (Wuhan) and (B) receptor binding domain (RBD) specific IgM, IgG1, and IgA1 binding levels were determined by Luminex in children receiving either 10 μg of BNT162b2 (ages 5-11 years old, yellow) or 30 μg BNT162b2 (ages 12-16 years old, blue and 16+ years old, red) before (V0_10μg (5-11y)_: 32; V0_30μg (12-16y)_: 29, V0_30μg (16+y)_: 7), after the first dose (V1_10μg (5-11y)_: 32; V1_30μg (12-16y)_: 27, V1_30μg (16+y)_: 8), or after the second dose (V2_10μg (5-11y)_: 30; V2_30μg (12-16y)_: 26, V2_30μg (16+y)_: 11). A two-way ANOVA (two-sided) was used to calculate statistically significant differences between the groups at each timepoint. Exact p-values for statistically significant differences after Benjamini-Hochberg correction for multiple testing are shown above the graph.

### Fc effector activity is robust but delayed in children 5-11 years old

Emerging data point to an important role for antibody Fc-effector functions in natural, vaccine induced, and therapeutic protection against COVID-19^11,18,19,21-23^. Specifically, in addition to neutralization, antibody recruit effector functions via interactions between their Fc domains and Fc receptors found on all immune cells^24^. Hence, we analyzed the Fc receptor (FcR) binding properties of SARS-CoV-2 Spike specific antibodies across the 3 immunized groups (**Figure 2A**). After the first dose, more robust cross-FcR binding profiles were observed in the adolescents and adults, both receiving the 30ug vaccine dose. Interestingly, adults raised a faster FcγR3a (p<0.001) and FcγR3b (p≤0.03) responses compared to adolescents who received the matching 30 μg dose. In contrast, after two doses adolescent Fc receptor binding was higher, albeit not significantly, compared to adults across nearly all FcγRs. Significantly higher binding to the IgG receptors FcγR2a (p_V1_=0.001; p_V2_=0.005), FcγR2b (p_V1_=0.001; p_V2_=0.002), FcγR3a (p_V1_<0.001; p_V2_=0.01) and FcγR3b (p_V1_=0.002 p_V2_<0.001) was seen in the adolescent group compared the younger 5-11 year old group after first and second vaccination. However, 5-11 year olds mounted similar levels of FcR binding to adults across FcγRs, albeit the responses were more variable in young children. Finally, low binding of IgA to its FcR receptor, FcαR, were noted across all 3 groups. Thus, while adolescents clearly generate more Spike-specific FcR-binding antibodies with the capacity to bind to FcRs more effectively than adults at the same dose. Conversely, 5-11 years olds induced comparable levels of FcR binding antibodies, at a lower antibody titer (**Figure 1**) compared to adults, suggesting that children generate qualitatively superior functional antibodies compared to adults.

**Figure 2.**
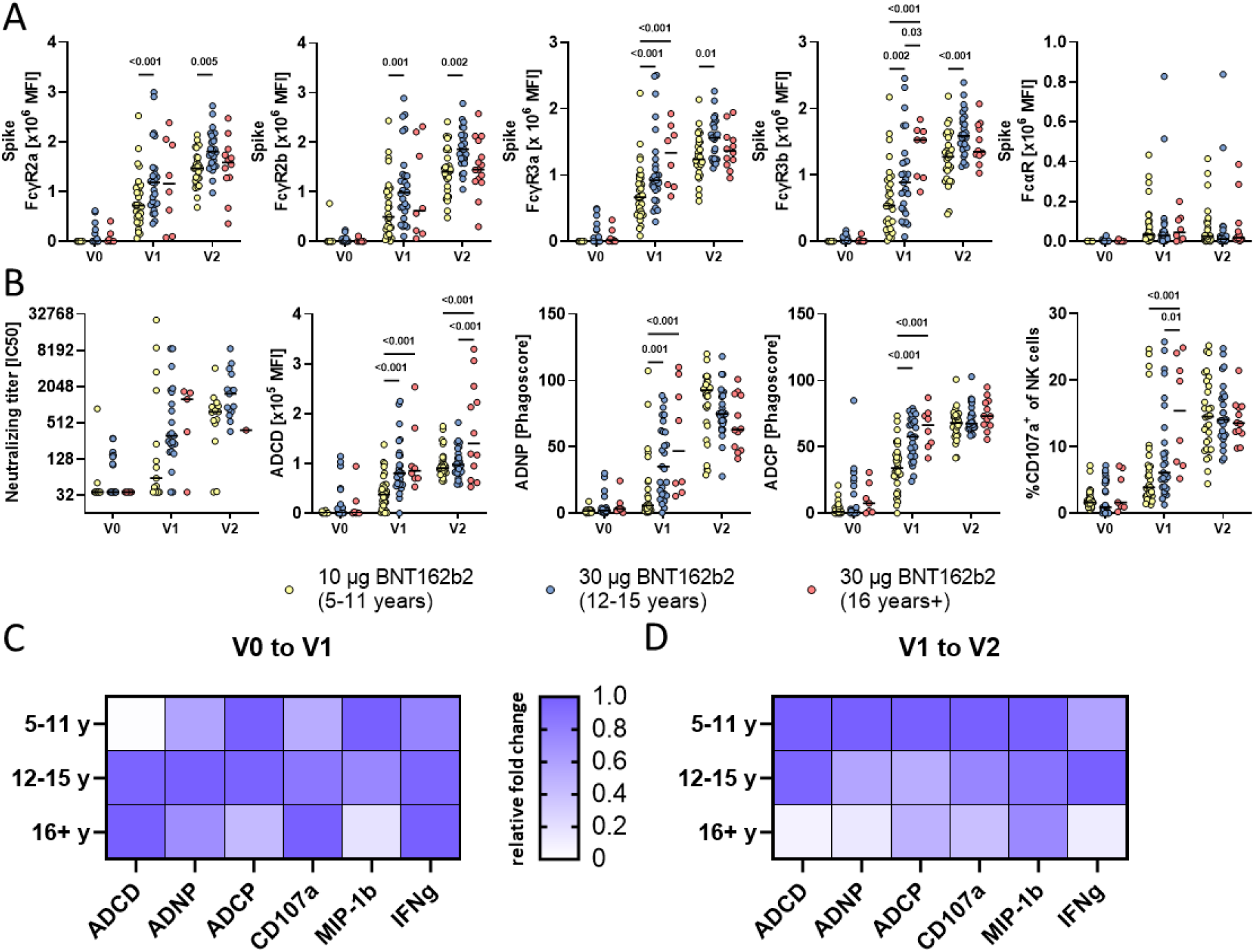
Vaccination with BNT162b2 mRNA in children induces higher FcγR, complement deposition, phagocytic activity, and NK cell activation. (A) Binding of SARS-CoV-2 spike specific antibodies to FcγR2a, FcγR2b, FcγR3a, FcγR3b and FcαR determined by Luminex in children receiving 10μg of BNT162b2 (ages 5-11 years old, yellow) or 30 μg BNT162b2 (ages 12-16 years old, blue and 16+ years old, red) before (V0_10μg (5-11y)_: 32; V0_30μg (12-16y)_: 29, V0_30μg (16+y)_: 7), after the first dose (V1_10μg (5-11y)_: 32; V1_30μg (12-16y)_: 27, V1_30μg (16+y)_: 8), or after the second dose (V2_10μg (5-11y)_: 30; V2_30μg (12-16y)_: 26, V2_30μg (16+y)_: 11). (B) The ability of SARS-CoV-2 spike specific antibody Fc to induce neutralization, complement deposition (ADCD), neutrophil phagocytosis (ADNP), monocyte phagocytosis (ADCP) and NK cell activation by the frequency degranulated CD107+ NK cells. A two-way ANOVA (two-sided) was used to calculate statistically significant differences between the groups at each timepoint. Exact p-values for statistically significant differences after Benjamini-Hochberg correction for multiple testing are shown above the graph). (C-D) Heatmaps show the relative fold changes of Fc mediated function before to after the first dose (V0 to V1)(C) or from the first to the second dose (V1 to V2)(D).

While antibody titers and FcR binding levels were more variable in younger children, receiving the lower vaccine dose, we next analyze whether antibody effector profiles differed across the groups (**Figure 2B**). While young children clearly required 2 doses of the vaccine to induce robust neutralizing antibody levels, both groups of children raised comparable antibody levels after the 2^nd^ dose, arguing that young children have the capacity to raise nearly equivalent neutralizing antibody titers to adolescents who received a higher dose of the vaccine. Of note, comparable to binding titers, some children and adolescent had pre-existing neutralizing titers potentially from cross-reactive antibodies. Conversely, more differences were noted across the 3 groups with respect to antibody effector functions. While all Fc effector functions were detected in all groups after the first dose and further enhanced after the second dose (**Figure 2B**), the kinetics of evolution of these responses was delayed in the 5-11 year old children. Specifically, adolescents and adults generated comparable antibody dependent complement depositing (ADCD), antibody dependent neutrophil phagocytic antibodies (ADNP) and antibody dependent cellular monocyte phagocytosis (ADCP), adults generated superior NK cell activating antibodies (CD107a) after a single dose. Yet, after the second dose, low-dose vaccinated children’s antibodies were superior at inducing ADNP, and induced comparable ADCP and ADNKA compared to high-dose adolescents and adults. When comparing the relative fold-induction after the first dose (V0 to V1) Fc effector functions were relatively uniformly enhanced across the groups (**Figure 2C**). Interestingly, in contrast, fold-changes after the second dose for all Fc-mediated functions (except IFNγ expression of NK cells which was higher in adolescent) were considerably higher in younger children compared to the adolescent group, and adults responded the weakest to the second dose. Given that the magnitude of these functional properties was not significantly different across the groups after the second dose, these data further highlight the importance of the second dose in children, whose antibodies, despite significantly lower subclass/isotype titers, were equally able to induce robust SARS-CoV-2 specific effector functions exclusively. Thus, young children experience a unique functional maturation, even at a lower vaccine dose, that may be attributable to their younger, more flexible immune response.

### Humoral immune responses are distinct across the groups

Given the differences in titers but less apparent differences in antibody function across the groups, we next aimed to define whether a multivariate profile may provide enhanced insights in the overall immunologic differences across the groups. Specifically, focusing on peak immunogenicity, after the complete two dose series, a supervised Least Absolute Shrinkage and Selection Operator (LASSO) was initially used to define the minimal features that could separate the groups, to avoid overfitting, and then the data were visualized using a Partial Least Square Discriminant Analysis (PLS-DA)(**Figure 3**). Vaccine induced antibody Fc-profiles diverged across the children (10μg dose) and adolescents (30μg dose) (**Figure 3A**), marked by elevated levels of Spike-specific IgM, IgA2, IgG1 and IgG3 titers as well as FcγR2a binding and IFNγ secretion by NK cells in adolescents. Conversely, vaccine-specific ADNP and NK cell MIP-1β secretion were selectively enriched in children. Similarly, comparison of children and adults separated more distinctly in the PLS-DA, with NK cell features including MIP-1β and IFNγ production enriched in children and IgG1, IgA and IgM titers, FcγR3b binding, ADCD, and ADCP enriched in adults (**Figure 3B**). Interestingly, even adolescent and adult responses differed, despite the matching vaccine dose, marked by age-dependent induction of higher vaccine-specific IgM, IgA and ADCD in adults, but increased binding to FcγR2a and monocyte phagocytosis (ADCP) in adolescents (**Figure 3C**). These data clearly illustrated the induction of higher antibody titers in adolescents and adults, but the unique expansion of functional responses in children.

**Figure 3.**
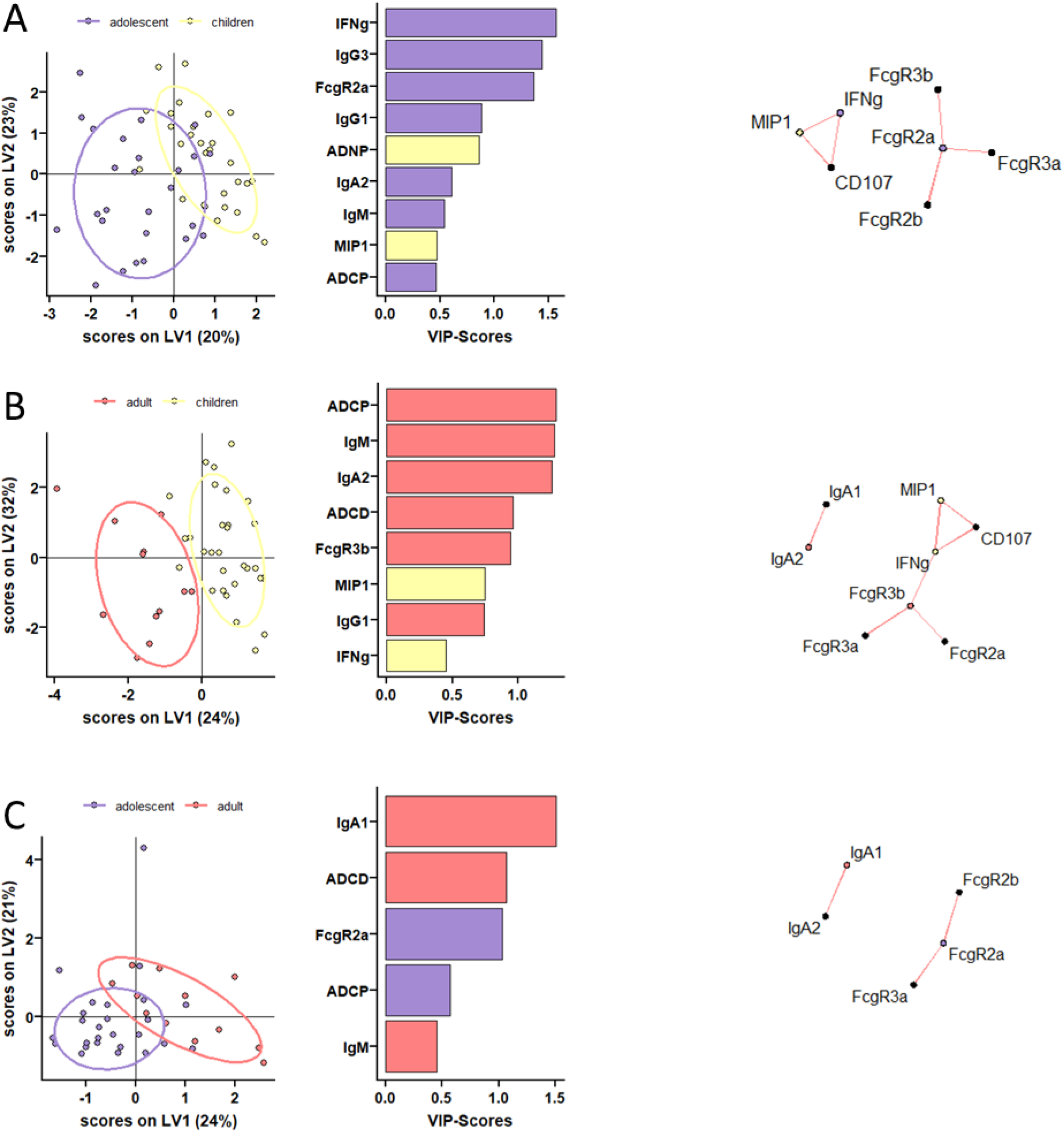
Distinct humoral profiles between BNT162b2 mRNA vaccinated pediatric children, adolescent children, and adults against SARS-CoV-2 wild-type spike. (A-C) A machine learning model was build using LASSO selected SARS-CoV-2 wild-type spike specific features at V2. Vaccine response in adolescent children (purple) and pediatric children (yellow) given 10 μg BNT162b2 (A), in adults (red) and pediatric children (yellow) (B), or in adults (red) and adolescent children (purple) given 30 μg BNT162b2 (C) were compared. Separation of the groups in the PLS-DA is shown in the right panel. Variable importance (VIP) score of LASSO selected features is shown in the bar graph and features are colored coded by the group they were enriched in. The network plots (right panel) show significant (p<0.05) Spearman correlations (|r|>0.7, only positive correlations were observed) to other (non-selected) features.

### Variable responses to variants of concern after BNT162b2 vaccination

The emergence of novel variants of concern (VOCs) like the most recent Omicron variant has led to continued waves of SARS-CoV-2 infections and breakthroughs globally. Dramatic antigenic changes particularly in the Omicron Spike Receptor Binding Domain – important for viral entry – led to near complete loss of vaccine-induced neutralizing antibody titers^25,26^. However, antibodies are still able to sense other parts of Spike VOCs, elicit Fc effector functions, and thereby potentially mediate protection from severe disease. However, whether vaccination in children leads to recognition of VOCs is uncler.

Thus, we next analyzed the specific humoral response to different VOCs, including alpha (B.1.1.7), beta (B.1.351), gamma (P.1), delta (B1.617.2) and omicron (B.1.1.529) at peak immunogenicity (**Figure 4**). Despite the lower antibody titers in children, all 3 groups exhibited remarkably stable IgA and IgG binding across VOCs (**Figure 4A**). In contrast, IgM binding antibodies was only decreased against omicron, pointing an isotype specific compromised response to this more mutated VOC, however all 3 groups clearly adapted their more mature IgA and IgG responses to compensate for this deficit in IgM immunity. Unexpectedly, IgM reactivity to the Beta variant was improved in children and adolescents compared to the D614G response pointing to age dependent differences in the evolution of breadth of binding across VOCs. Moreover, fold changes of binding over D614G across isotype and FcR binding titers to different VOC full Spike and RBDs highlighted consistent patterns of breadth of binding across VOCs across the age groups (**Figure 4B**), with the exception of IgM binding in children and adolescents that was superior across all VOCs, except Omicron. These data point to a unique flexibility in cross-reactivity in IgM in children. Given the robust opsonophagocytic and complement fixing function of IgM, these data point to a unique capability build into to youth that may help provide some level of persistent broad recognition of VOCs following vaccination.

**Figure 4.**
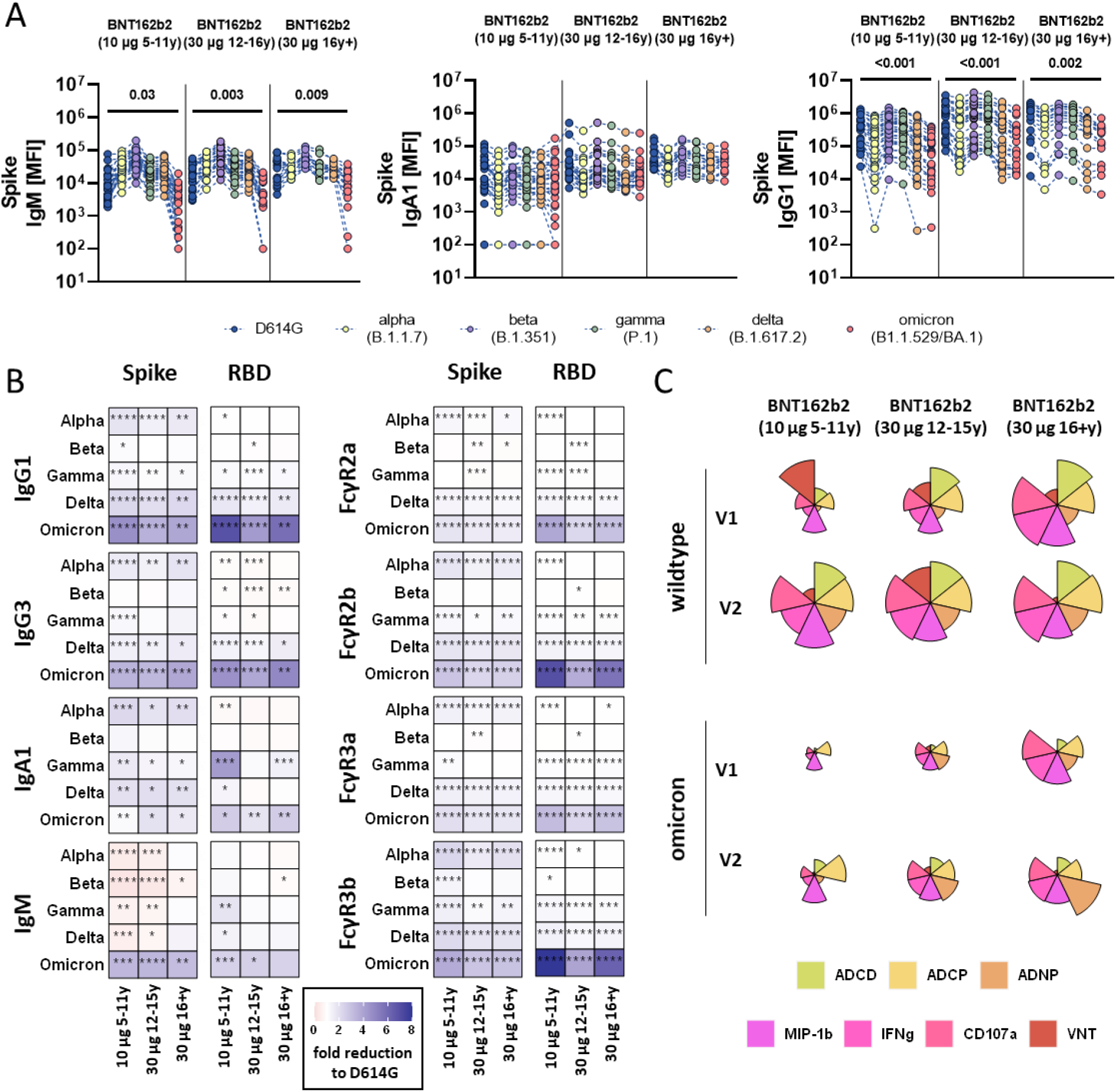
SARS-CoV-2 variants of concern specific humoral immune responses after vaccination with BNT162b2. (A) Vaccine induced IgM, IgA1, and IgG1 response to D614G (wild-type; blue), alpha (B.1.1.7, yellow), beta (B.1.1.7, purple), gamma (P.1, green), delta (B.1.617.2, orange), and omicron (B1.1.529/BA.1, red) to the full Spike in children receiving 10μg of BNT162b2 (ages 5-11 years old, n =) or adolescent receiving 30 μg BNT162b2 (ages 12-15 years old, n=) or adults (16+ years old, n=) at V2. A two-sided Kruskal-Wallis test with Benjamini-Hochberg correction for multiple testing was performed to compare D614G and omicron specific antibody titers. P-values for significant different comparisons are shown above the dataset. B) Heatmaps show the fold change (red=increase, blue= decrease) for the different VOCs compared to the original D614G variant for Spike and RBD specific IgG1, Ig3, IgA1 and IgM titers or binding to FcγR2a, FcγR2b, FcγR3a, FcγR3b. C) Flower plots summarize ADCD, ADCP, ADNP, ADNKA (CD107a, IFNγ, MIP-1β) and neutralization (VNT) at V1 and V2 against D614G (upper panel) or omicron (lower panel) Spike in 10μg of BNT162b2 (ages 5-11 years old) or adolescent receiving 30 μg BNT162b2 (ages 12-15 years old) or adults (16+ years old) at V2. Each petal represents a specific function (compare color key) and the length of the petal corresponds of the intensity of Z-scored and normalized data. Asterisks in B) indicate significant differences of the respective variant in a paired two sided Wilcoxon rank test. P-values were corrected for multiple testing using Benjamini-Hochberg correction. *:p<0.05, **:p<0.01,***:p<0.001, ****:p<0.0001.

While antibody binding to most VOC Spikes was largely preserved, there was an apparent reduction for the Omicron variant recognition across all ages. We therefore profiled the ability of vaccine-induced antibodies to elicit neutralizing and Fc-mediated effector functions to Omicron (**Figure 4C**). As expected, Omicron specific antibody effector functions were detectable but reduced when compared to D614G Spike-specific responses after first and second dose of BNT162b2. Children receiving the 10 μg dose, showed lower Omicron reactivity at both timepoints compared to adult-dosed adolescent or adults. Interestingly, while the Omicron effector profiles expanded from the first to the second dose in the 10 μg dosed children and 30 μg dosed adolescents, ADNKA and ADCP to omicron were reduced from the first to the second dose in adults. Collectively, our data indicate that while adults produce sufficient effector responses after the first vaccine dose, adult immunity becomes narrower towards the vaccine insert antigen after the second dose. In contrast, children and adolescents may not be fully primed after the first dose, but develop a broad and highly functional vaccine response after the full vaccine series.

## Discussion

Early in the pandemic, pediatric cases accounted for approximately 5% of the total COVID-19 cases in the US. Reasonably, initial vaccine campaigns focused on the protection of adults. Since then, pediatric cases of SARS-CoV-2 have exceeded 13 million with over 42,000 hospitalizations^3^, resulting in the emergency use authorization for mRNA vaccines for children 5 years of age and older. However, due to the delayed vaccine rollout and increased vaccine hesitancy by parents, vaccination rates in children still lag behind. Together with the occurrence of novel variants like omicron accompanied by relaxed masking and social distance measurements, cases of COVID-19 have soared with up to 20% of the total SARS-CoV-2 infections occurring in children during the recent omicron wave^3^. In addition to acute infection, children are at special risk of developing a severe multi-inflammatory syndrome (MIS-C) weeks after asymptomatic SARS-CoV-2 exposure. Importantly, data indicates that 30 μg BNT162b2 not only protect adolescents (12-18 years) from severe COVID-19 but also MIS-C^27^, underscoring the importance and effectiveness of vaccination across the ages.

As innate and adaptive immune systems mature during the first decades of life, it was not clear how the naïve, juvenile immune system would respond to novel mRNA vaccine technology. Furthermore, due to safety and tolerance concerns, doses were adjusted for the younger 5-11 year old age group. Our data point to a more variable immune response in children 5-11 years of age receiving a three-fold lower dose BNT162b2 compared to adolescents and adults. Analysis of a dose-down study of the Moderna mRNA-1273 vaccine suggests that children have an attenuated immune response after a two-fold lower-dose, though, age specific differences may have also accounted for this^23^. Here, we noted a selective humoral deficiency in isotype and Fc receptor binding titers after one dose of BNT162b2 in children and adolescent compared to adults. Yet, humoral immunity expanded robustly after the second dose, raising functions to levels comparable or superior to those observed in adults, with titers in the adolescent group exceeding those in adults. Moreover, after two doses, children had specifically elevated levels of opsonophagocytic and NK cell activating antibodies that have been previously associated with resolution of severe disease^28,29^. This enhanced functionality could be attributable to a variation in epitope selection or post-translational modification of the antibody Fc-domain that collectively could alter the quality of the pediatric immune response antibody quality. Furthermore, antibody avidity was higher in children and adolescents (**Fig S2**) pointing to qualitatively superior immune responsiveness to the response to mRNA vaccination in children and adolescents. Additionally, whether a matched adult dose of mRNA or expanded interval between vaccine doses could further expand and improve the 5-11 year old immune response remains unclear but warrants further investigation into dose dependent immune programming across the ages to guide future of mRNA vaccines.

Since the beginning of the SARS-CoV-2 pandemic, cross-reactive antibodies, and repetitive exposure of young individuals to other circulating coronaviruses have been discussed as potential contributors to shaping SARS-CoV2 immunity and providing protection against severe disease. While causal evidence is still missing, several studies have described the existence of pre-pandemic cross-reactive antibodies or T cell epitopes to SARS-CoV-2 that have appear to reduce disease severity ^30,31^. Moreover, other studies have argued that the sequence of VOC exposure can lead to original antigenic sin restricting the quality and breadth of the overall humoral immune response ^32,33^. Likewise, we noted pre-existing SARS-CoV-2 Spike specific antibodies able to mediate neutralization and elicit Fc effector functions, in the absence of evidence of previous infection (**Fig S2**). Along these lines, immune responses to common coronaviruses, such as the Beta-coronaviruses HKU1 and OC43, were correlated to antibody binding to SARS-CoV-2 Spike across populations (**Fig S2**). Furthermore, BNT162b2 vaccination boosted pre-existing and elicited novel OC43 specific antibodies and was associated with post-vaccine titers across the groups, pointing to limited evidence of the original antigenic sin. However, whether vaccine titers limit the development of antibodies to more related SARS-CoV-2 VOCs (e.g., after natural infection or VOC specific vaccination) needs to be addressed in the future.

Here, we investigated vaccine response in children and adolescent after the currently recommended “real-world’ doses. While slight dose-dependent changes were observed in initial clinical trials in 5-11 year olds^34^, it remains elusive whether the observed differences here are due to differences in age, dose or both. We analyzed vaccine responses at the expected peak immunogenicity two weeks after the second dose, and it will be critical in the future once the samples become available to analyze durability of these response to guide additional booster recommendations as for the adult population. Furthermore, while distinct antibody functions and vaccine profiles have been linked to protection, it is unclear, whether the observed differences here translate to differences in disease attenuation, in particular against emerging VOCs. While we observed some association of pre-existing and vaccine induced SARS-CoV-2 antibodies to other beta-coronavirus titers, our sample size was too small to assess whether distinct age-specific differences in the exposure to theses circulating “common-cold” coronaviruses could explain differences in the vaccine response.

Vaccination against SARS-CoV-2 has been proven safe and effective. Yet, the occurrence of novel variants of concerns, potentially fueled by vaccine hesitancy and delayed rollout in some parts of the world, have challenged vaccine-induced immunity. However, antibody binding to most VOCs was largely preserved after mRNA vaccination. Yet, the dramatic antigenic changes in the RBD of the recent omicron VOC, have led to substantial loss of neutralizing activity of vaccine-induced antibodies and surge in vaccine breakthrough cases. Vaccination still protects from severe disease and hospitalization indicating that other vaccine-induced immune mechanism are more conserved. Here, we observed a pronounced loss of omicron RBD specific antibodies tracking with loss of neutralization. Still, other parts of the Omicron Spike were readily sensed and antibodies able to mediate additional Fc effector functions potentially mediating an additional layer of vaccine-induced protection. Fc effector functions to Omicron were readily detected in adults (16+). In contrast, children elicited comparable cross-reactivity, albeit at a lower magnitude, and reduced omicron antibody functionality, likely attributable to the lower vaccine dose. While these responses likely continue to recognize potential infections and limit disease, augmentation of these responses with boosting or adjusted dosing may provide a more robust barrier to control the virus within this unique population. Along these lines, stability of the more variable response in young children needs to be critically monitored to guide booster immunization guidelines and to shape future mRNA vaccine strategies in children.

## Data Availability

All data produced in the present work are contained in the manuscript.

## Contributions

Y.C.B., J.W.C, and J.K. performed the serological experiments. K.J.D, M.L.S. and A.B.B. performed the neutralization assay. Y.C.B., L.M.Y. and G.A. analyzed and interpreted the data. M.D.B., J.P.D., and L.M.Y. supervised and managed the sample collection. G.A. supervised the project. Y.C.B., L.M.Y. and G.A. drafted the manuscript. All authors critically reviewed the manuscript.

## Acknowledgment

We thank Nancy Zimmerman, Mark and Lisa Schwartz, an anonymous donor (financial support), Terry and Susan Ragon, and the SAMANA Kay MGH Research Scholars award for their support. We acknowledge support from the Ragon Institute of MGH, MIT, and Harvard, the Massachusetts Consortium on Pathogen Readiness (MassCPR), the NIH (3R37AI080289-11S1, R01AI146785, U19AI42790-01, U19AI135995-02, U19AI42790-01, 1U01CA260476 – 01, CIVIC75N93019C00052, 5K08HL143183, 1R01HD100022-01 and 3R01HD100022-02S2), the Gates Foundation Global Health Vaccine Accelerator Platform funding (OPP1146996 and INV-001650), and the Musk Foundation.

## Methods and Materials

### Sample Collection

Pediatric samples were collected from children vaccinated with either 10 μg BNT162b2 (ages 5-11 years old) or 30 μg BNT162b2 (ages 12-18 years old) as a part of FDA’s emergency use authorization of the BNT162b2 vaccine for these age groups. Sera were collected prior to vaccination (V0), before the second dose approximately 3 weeks after the first dose of Pfizer-BioNTech mRNA vaccination (V1), and/or two to four weeks after the second dose (V2). All individuals or their legal guardian gave informed consent prior to enrollment. This study was overseen and approved by the MassGeneralBrigham Institutional Review Board (#2020P000955).

### Cell Lines and Primary Cells

THP-1 cells, a human leukemia monocyte cell line, were maintained in RPMI-1640 Medium (Sigma-Aldrich) with 10% fetal bovine serum (FBS, Sigma-Aldrich), 1% L-glutamine (Corning), 2% of 5000 IU/mL of Penicillin and 5,000 ug/ml of Streptomycin Solution (Pen-Strep, Corning), 1% of 1M HEPES (Corning), and 5mM 2-Mercaptoethanol (Gibco). Cells were incubated at 37°C with 5% CO_2_.

Primary neutrophils for antibody-dependent neutrophil phagocytosis (ADNP) assays were isolated from whole blood from healthy donors using Ammonium-Chloride-Potassium (ACK) Lysing Buffer (Quality Biological) followed by centrifugation and multiple wash steps prior to assay usage.

Primary natural killer cells for the antibody-dependent natural killer activation (ADNKA) assays were isolated from buffy coats from health donors using the RosetteSep NK cell enrichment kit (STEMCELL Technologies). Cells were incubated overnight at 37°C with 5% CO_2_ in RPMI-1640 Medium (Sigma-Aldrich) with 10% FBS (Sigma-Aldrich), 1% L-glutamine (Corning), 2% of Pen-Strep (Corning), and 1% of 1M HEPES (Corning) prior to assay usage.

Human embryonic kidney (HEK) 293T cells expressing ACE2 for neutralization assays were maintained in Dulbecco’s Modified Eagle’s Medium (DMEM, Corning) with 10% FBS (VWR) and 1% Pen-Strep (Corning) and incubated in 37°C with 5% CO_2_.

### Antigens and Biotinylation

Antigens were biotinylated using the NHS-Sulfo-LC-LC kit following manufacturer’s instructions (ThermoFisher). Excess biotin was removed via size exclusion chromatography by using Zeba-Spin desalting columns (7 kDa cutoff, ThermoFisher).

### Antibody Isotype and Fc Receptor Binding

Antigen-specific antibody isotype, subclass, and Fc receptor binding profiles were analyzed using a custom multiplex Luminex assay as previously described ^35^. The antigens were then coupled directly onto the Luminex microspheres (Luminex Corp) via carboxy coupling. Coupled beads were incubated with diluted plasma samples overnight at 4°C. Following overnight incubation, non-specific antibodies were washed off and the immune complexes were incubated with Ig isotypes or subclasses with a 1:100 diluted PE-conjugated secondary antibody for IgG1 (clone: HP6001), IgG2 (clone: 31-7-4), IgG3 (clone: HP6050), IgG4 (clone: HP6025), IgM (clone: SA-DA4), IgA1 (clone: B3506B4), or IgA2 (clone: A9604D2) (all Southern Biotech). For the FcγR binding, the secondary probe is a PE-streptavidin (Agilent Technologies) coupled recombinant and biotinylated human FcγR protein. Beads with secondary antibodies were incubated on a shaker at room temperature for 1 hour. Following incubation, excessive antibodies were washed off and relative antigen-specific antibody levels were determined on an iQue analyzer (Intellicyt).

### Antibody-Dependent Complement Deposition (ADCD) Assays

ADCD assays were performed as previously described ^36^. Biotinylated antigens were coupled to FluoSphere NeutrAvidin beads (ThermoFisher). Immune complexes were formed by incubating of antigen coupled beads with 10 ul of 1:10 diluted plasma samples at 37°C for 2 hours. Following incubation, non-specific antibodies were washed off followed by incubation of the immune complex with guinea pig complement in GVB++ buffer (Boston BioProducts) at 37°C for 20 minutes. To stop the complement reaction, 15 mM EDTA (Corning) in PBS was added to the immune complex. C3 deposited on beads were stained with anti-guinea pig C3-FITC antibody (MP Biomedicals, 1:100, polyclonal) and analyzed on an iQue analyzer (Intellicyt).

### Antibody-Dependent Neutrophil Phagocytosis (ADNP) Assays

ADNP assays were performed as previously described ^37^. Biotinylated antigens were coupled to FluoSphere NeutrAvidin beads (ThermoFisher). Immune complexes were formed by incubating of antigen coupled beads with 10 ul of 1:50 diluted plasma samples at 37°C for 2 hours. Primary neutrophils were isolated as previously mentioned and incubated with washed immune complexes at 37°C for 1 hour. Following incubation, neutrophils were stained for surface CD66b (Biolegend, 1:100, clone: G10F5) expression and fixed with 4% para-formaldehyde. Analysis was done on an iQue analyzer (Intellicyt).

### Antibody-Dependent Cell Phagocytosis (ADCP) Assays

ADCP assays were performed as previously described ^38^. Biotinylated antigens were coupled to FluoSphere NeutrAvidin beads (ThermoFisher). Immune complexes were formed by incubating 10 ul of antigen coupled beads with 10 ul of 1:100 diluted plasma samples at 37°C for 2 hours. THP-1 monocytes were then added to the beads then incubation at 37°C for 16 hours. Following incubation, cells were fixed with 4% para-formaldehyde and analyzed on an iQue analyzer (Intellicyt).

### Antibody-Dependent NK Activation (ADNKA) Assays

MaxiSporp ELISA plates (ThermoFisher) were coated with either 1ug/ml SARS-CoV-2 wt spike or omicron variant spike at room temperature for 2 hours followed by blocking with 5% BSA (Sigma-Aldrich). 50 ul of 1:50 diluted plasma samples were added to the wells and incubated overnight at 4°C. Primary NK cells were prepared as described above. Cells were added to the washed ELISA plate and incubated with anti-human CD107a (BD, 1:40, clone: H4A3), brefeldin A (Sigma-Aldrich) and monensin (BD) at 37°C for 5 hours. Following incubation, cells were surface stained for CD56 (BD, 1:200, clone: B159) and CD3 (BD, 1:200, clone: UCHT1). The NK cells were then washed, fixed, and permeabilized using the Fix & Perm Cell Permeabilization Kit (ThermoFisher). Cells were then stained for intracellular markers MIP1β (BD, 1:50, clone: D21-1351) and IFNγ (BD, 1:17, clone: B27). NK cell populations were defined to be CD3-CD56+. The frequency of degranulated cells marked by CD107a and the expression of intracellular INFγ and MIP-1β were determined on an iQue analyzer (Intellicyt).

### Virus Neutralization

Virus neutralization was performed as previously described ^39^. Three-fold serial dilutions of plasma samples starting at 1:12 or 1:30 were performed before adding pseudovirus expressing either SARS-CoV-2 wild-type spike or omicron variant spike to HEK293T expressing ACE-2 cells for 1 hour.

Pseudoviruses were produced by PEI transfection of lentiviral vector with CMV-Luciferase-IRES-ZsGreen, lentiviral helper plasmids, and either SARS-CoV-2 wt spike or omicron variant spike expression plasmid. Pseudovirus neutralization titers (pNT50) values were calculated by taking the inverse of 50% inhibitory concentration value for all samples with a pseudovirus neutralization value of 80% or higher at the highest concentration of serum.

### Data Analysis and Statistics

Data analysis was performed on GraphPad Prism (v.9.3) and RStudio (v.1.3). A two-way ANOVA test in GraphPad Prism was used to determine statistically significant differences between the groups at each timepoint. Flower plots were generated with the ggplot package in R using Z-scored data. Multivariate classification models were built to discriminate between humoral profiles of vaccination arms. Feature selection was performed using the least absolute shrinkage and selection operator (LASSO) and classification and visualization were performed using partial least square discriminant analysis (PLS-DA). These analyses were performed using R package “ropls” version 1.20.0 ^40^ and “glmnet” version 4.0.2^41^. Co-correlate networks were built using Spearman method followed by Benjamini-Hochberg correction and the co-correlate network was generated using R package “network” version 1.16.0^42^.

