## Supplementary material for "BNT162b2 induces robust cross-variant SARS-CoV-2 immunity in children": Suppl

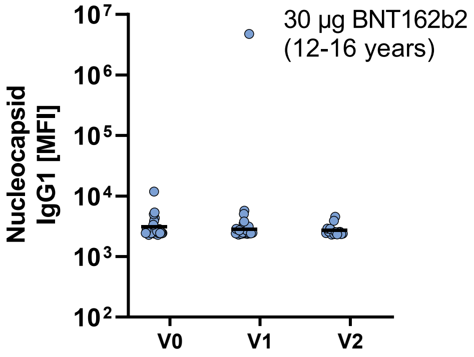


**Fig S1: Nucleocapsid specific antibody titer show no signs of previous infection.** SARS-CoV-2 specific Nucleocapsid IgG1 titers were determined in 12-15 year old individuals before (V0, n=29) and after first (V1, n=29) and second (V2, n=15) dose 30 µg BNT162b2 by Luminex.

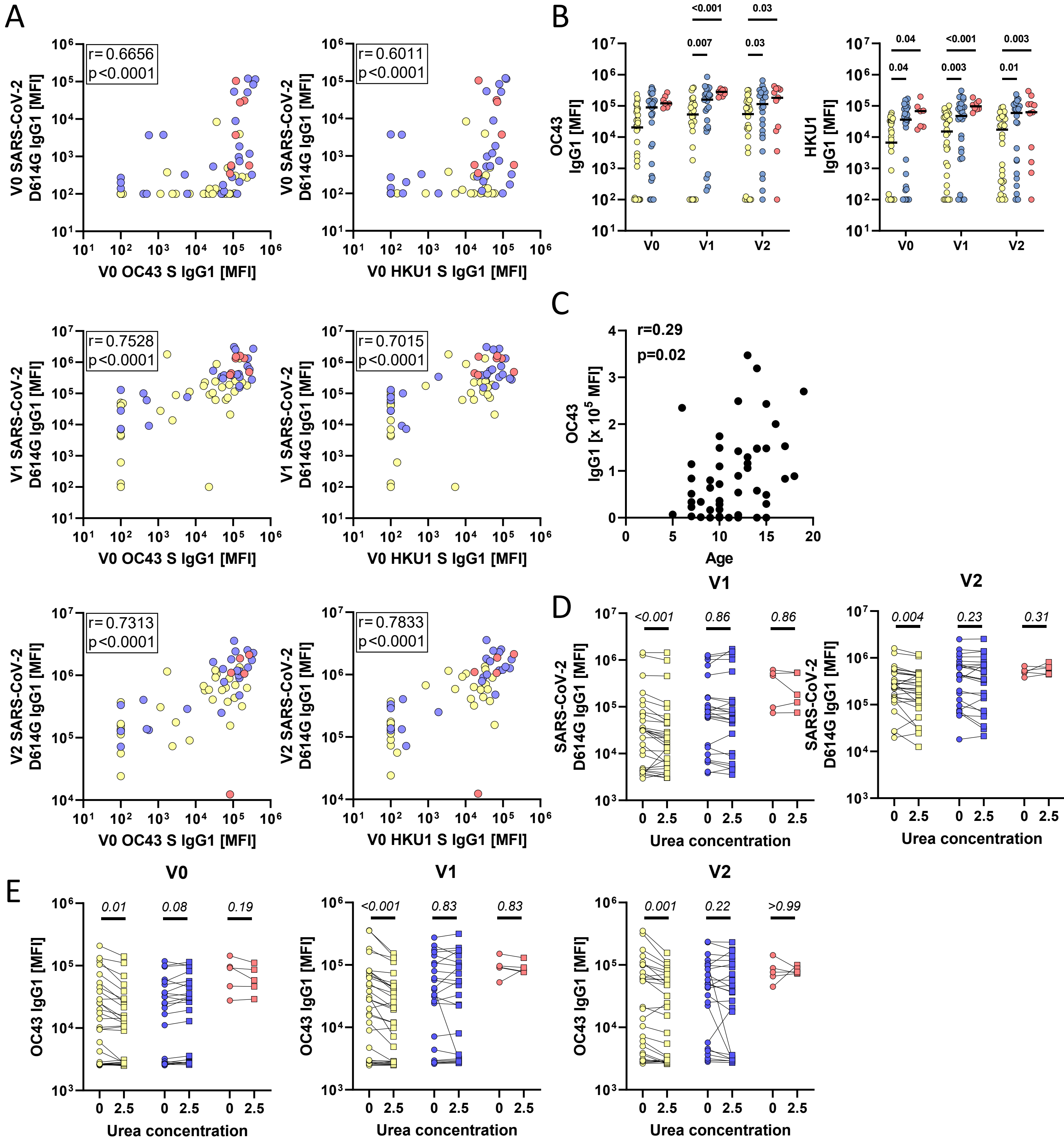


**Fig S2: Relationship of SARS-CoV-2 and beta-coronavirus titer.** A) SARS-CoV-2 Spike specific IgG1 titers at V0 (top), V1 (middle) or V2 (bottom) were plotted against OC43 or HKU1 specific IgG1 titers at V0. Spearman correlations between SAR-CoV-2 Spike and respective beta-coronavirus were calculated and coefficient (r) and p-value indicated in the plot. B) OC43 and HKU1 specific IgG1 titers Luminex in children receiving either 10 µg of BNT162b2 (ages 5-11 years old, yellow) or 30 µg BNT162b2 (ages 12-16 years old, blue and 16+ years old, red) before ($V0_{10\mu g\,(5\text{-}11y)}$: 32; $V0_{30\mu g\,(12\text{-}16y)}$: 29, $V0_{30\mu g\,(16+y)}$: 7), after the first dose ($V1_{10\mu g\,(5\text{-}11y)}$: 32; $V1_{30\mu g\,(12\text{-}16y)}$: 27, $V1_{30\mu g\,(16+y)}$: 8), or after the second dose ($V2_{10\mu g\,(5\text{-}11y)}$: 30; $V2_{30\mu g\,(12\text{-}16y)}$: 26, $V2_{30\mu g\,(16+y)}$: 11). C) Correlation of OC43 specific titers with age. Spearman correlation was calculated and coefficient (r) and p-value indicated in the plot. D-E) Avidity of SARS-CoV-2 (D) or OC43 (E) antibodies was assessed by washing Luminex beads after sample incubation with 2.5 M urea and compared to the native (0M urea) value ($V0_{10\mu g\,(5\text{-}11y)}$: 30; $V0_{30\mu g\,(12\text{-}16y)}$: 23, $V0_{30\mu g\,(16+y)}$: 5, $V1_{10\mu g\,(5\text{-}11y)}$: 30; $V1_{30\mu g\,(12\text{-}16y)}$: 24, $V1_{30\mu g\,(16+y)}$: 5, $V2_{10\mu g\,(5\text{-}11y)}$: 28; $V2_{30\mu g\,(12\text{-}16y)}$: 24, $V2_{30\mu g\,(16+y)}$: 5).
